# Health Status of Puerto Rican Youth in the Island: A Survey of Key Cardiometabolic and Mental Health Aspects

**DOI:** 10.64898/2026.03.21.26348983

**Authors:** Marisela Irizarry-Pérez, Breanna Beaumont, Augusto Enrique Caballero, Edmarie Guzmán-Vélez

## Abstract

**Background:** Puerto Rican young adults face elevated cardiometabolic, mental health, and healthcare access risks, yet this population remains understudied. We characterized cardiometabolic health, psychosocial well-being, and health-related behaviors among Puerto Rican young adults.

**Methods:** A cross-sectional survey conducted from 2022 to 2025 assessed self-reported cardiometabolic risk factors, mental health, loneliness, healthcare access and utilization, and attitudes toward health-promoting behaviors among Puerto Rican adults aged 21-35 years. We calculated prevalence estimates and used chi-square tests to examine differences by sex and age group.

**Results:** Of 2,783 participants (mean age 27.3; 72.7% female; 59.9% with >4 years of college), over 40% reported >6 days of poor mental health and 20.4% poor physical health in the past month. Most had health insurance (92.2%), yet over one-third faced financial barriers to care and lacked a primary care provider (PCP). Influenza vaccination was low (30.4%), while HPV vaccination (54.7%) and HIV testing (70.7%) were higher. Over half were overweight or obese (57.2%), 14.7% reported elevated blood glucose, and 10.9% hypertension. Females reported poorer mental health but greater preventive care engagement; males were more likely to lack a PCP and report hypertension and tobacco use. Younger adults reported worse mental health and higher loneliness.

**Conclusions:** Puerto Rican young adults carry a high burden of cardiometabolic risk, mental health challenges, and preventive care gaps. Further research should identify effective strategies to increase preventive care engagement and the adoption of healthy behaviors among this group, thereby reducing the chronic disease burden in later adulthood.

**KEY MESSAGES:** *What is already known on this topic:* Puerto Ricans experience a disproportionate burden of cardiometabolic and mental health conditions, but research has largely overlooked young adults, who came of age through economic crisis, natural disasters, and the COVID-19 pandemic.

*What this study adds:* This is the first post-pandemic, multi-domain study of Puerto Rican young adults, showing high rates of obesity, hypertension, poor mental health, and loneliness alongside major preventive care gaps, despite strong personal motivation toward healthy behaviors.

*How this study might affect research, practice or policy:* These findings support integrated interventions, expanded affordable preventive services, and community- and university-based screening to close the gap between young adults’ health motivation and their actual access to care.

## INTRODUCTION

The Hispanic/Latino population bears a disproportionate burden of chronic disease and associated risk factors relative to other racial and ethnic groups in the United States (US)^1,2^. Within this population, Puerto Ricans exhibit some of the poorest cardiovascular health profiles, with higher prevalences of obesity, diabetes, hypercholesterolemia, and hypertension than the US national population and most other Hispanic/Latino groups^2–4^, alongside less favorable lifestyle patterns including tobacco use, poor diet and sleep quality, and lower levels of physical activity than other Hispanic/Latino groups^5,6^. Critically, these disparities are not confined to older age, as young adult Puerto Ricans also exhibit suboptimal cardiovascular health, suggesting risk is established early and compounds over the life course^5–7^. These findings are especially concerning since poor cardiovascular health in young adulthood is a strong predictor of later cardiovascular disease, cancer, and dementia^8–10^.

Young adults are also among the least likely to have health insurance or to engage with primary care, factors essential for early disease detection and prevention^11^. Even when healthcare is accessed, nearly 70% of visits do not include preventive counseling and screening rates for conditions central to this age group, such as sexually transmitted infections, obesity, and injury risk, remain low^12^. Compounding these gaps in healthcare, risk-taking behaviors that contribute to long-term morbidity and mortality often emerge or peak during young adulthood, including tobacco use, binge drinking, high-risk sexual activity, physical inactivity, and poor dietary habits.^4,6^ Additionally, young adults are particularly vulnerable to mental health challenges, which have worsened in the wake of the COVID-19 pandemic^13,14^. Collectively, these patterns position young adulthood as a critical and underutilized window for intervention to prevent the development of chronic disease.

These risk patterns reflect the accumulation of socioeconomic disadvantage, chronic stress, and structural barriers to care^2,15,16^. Puerto Ricans on the island face lower household income and higher unemployment than their mainland counterparts despite relatively high educational attainment^17^, and receive disproportionately less Medicaid funding than US states, limiting preventive and specialty care^18^. Young adults are particularly underserved, aging out of pediatric systems without entering chronic disease prevention programs^19^. Compounding this, Puerto Rican young adults have experienced multiple large-scale stressors during critical developmental periods, prolonged economic crisis, Hurricane María, earthquakes, and COVID-19, exposures that may synergistically exacerbate risk across health domains and set the stage for decades of disease burden^15^.

Despite this well-documented burden, critical gaps persist. Most studies examine cardiometabolic risk, mental health, and healthcare access in isolation, leaving their co-occurrence poorly characterized. Population-based data specific to island-residing Puerto Ricans remain scarce, as the Hispanic/Latino health literature is dominated by mainland samples that do not reflect the island’s structurally distinct environment. Moreover, the majority of available evidence predates the COVID-19 pandemic and the sequence of compounding disasters Puerto Rico has experienced since 2017, limiting its relevance given substantial shifts in mental health, healthcare utilization, and lifestyle behaviors^12,13^. Collectively, these issues leave a critical gap in the evidence base needed to develop timely, contextually grounded prevention strategies for a population at elevated and compounding risk.

To address these gaps, this study provides the first contemporary, multi-domain assessment of cardiometabolic health, psychological well-being, healthcare access and utilization, and health-related behaviors among Puerto Rican young adults on the island, conducted in the post-pandemic era when cumulative stressor effects are most likely to be apparent. We further examine sociodemographic differences and perceptions of healthy behaviors and barriers to preventive care. We hypothesized that Puerto Rican young adults would exhibit a high and clustered burden of risk across cardiometabolic, behavioral, and mental health domains. Given documented sex differences in cardiometabolic risk factors and age-related patterns of mental health vulnerability, we further hypothesized that males would show less favorable cardiometabolic profiles and that younger individuals would report greater psychological distress.

## METHODS

### Study Design

We examined cross-sectional data from a community-based health education and prevention program for young adults aged 21–35 in Puerto Rico (Ponte Pa’ Ti), led by Varmed Management Group, LLC (VarMed). Questionnaires were distributed online between June 2022 and July 2025, promoted via social media with raffle-based incentives, including concert and sports event tickets, in both metropolitan and non-metropolitan areas t,o reduced geographic selection bias. Participants were eligible if they were aged 21–35, resided in Puerto Rico, and could respond in Spanish.

### Ethics Statement

The protocol was reviewed by the Advarra Institutional Review Board and determined not to constitute human subjects research. A fully de-identified dataset was shared by Varmed via a secure, encrypted platform in compliance with HIPAA regulations for secondary data analysis.

### Measures

Participants completed a self-administered Spanish-language questionnaire assessing demographic, biometric, healthcare, and lifestyle variables, incorporating validated items from the Behavioral Risk Factor Surveillance System (BRFSS)^20^. Demographic variables included age, sex assigned at birth, civil status, education level, area of residence (metropolitan or non-metropolitan), and employment status. Height and weight were used to calculate BMI, with participants categorized as underweight (<18.5), healthy weight (18.5–24.9), overweight (25–29.9), or obese (≥30).

Perceived general health was rated on a five-point scale (excellent to poor). Health-related quality of life was assessed by self-reported days of poor physical and mental health and activity-limiting health problems in the past 30 days.

Healthcare access and utilization variables included health insurance coverage, having a PCP, financial barriers to care, and time since last PCP and dental visit. Preventive care measures included influenza vaccination in the past year, HPV vaccination, HIV and STD testing, and blood glucose screening. Participants also reported prior diagnoses of hypertension or prediabetes. Females were additionally asked about Pap smear history.

Health attitudes were assessed using four Likert-scale items on the importance of physical activity, a balanced diet, mental health, and interest in developing healthier habits.

The second version of the survey, released in April 2024, included the 6-item De Jong Gierveld Loneliness Scale (DJGLS), a cross-culturally validated measure yielding emotional, social, and total loneliness scores, with higher scores indicating greater loneliness^21^. It also included items on cigarette and alcohol consumption frequency.

### Statistical analysis

Descriptive statistics were computed for all study variables. No formal a priori sample size calculation was performed. Data collection continued through the program’s standard enrollment period, yielding a final sample of 2,783 participants. Continuous variables are presented as means and standard deviations, and categorical variables as frequencies and percentages. Independent-samples *t* tests were used to compare continuous variables, and chi-square tests were used to examine differences in categorical variables between sex assigned at birth and age groups (21–25, 26–30, and 31–35 years). An analysis of variance (ANOVA) was conducted to compare differences in loneliness scores across age groups. Bonferroni corrections were applied to account for multiple comparisons. There was no missing data. Analyses were conducted in IBM SPSS (version 30), with significance set at P < 0.05.

## RESULTS

A total of 2,783 individuals completed the questionnaire. Of those, 2,337 completed the loneliness questionnaire. Most participants self-reported being female at birth, residing in a non-metropolitan area, being employed, being unmarried or not living with a partner, and not having children (**Table 1**). More than half of the participants fell in the overweight or obese categories (**Table 2)**. Overall, 92.2% rated their general health as at least very good. However, 20.4% and 40.5% reported experiencing six or more days of poor physical and poor mental health, respectively, in the past month, and 29.6% indicated that physical or mental health problems interfered with their usual activities for at least six days during that period. Nearly all participants reported having health insurance. Yet, over one-third indicated that they sometimes were unable to seek medical care due to financial constraints. Nearly one-third reported not having a PCP or other healthcare professional for routine care, and close to two-thirds indicated that it had been less than a year since their most recent routine visit to a PCP or dentist. Most individuals reported not receiving a flu shot in the past year. In contrast, most had been tested for HIV or other STDs, and slightly over half had received the HPV vaccine. Among females, approximately three-quarters reported having undergone a PAP smear.

**Table 1.** Sociodemographic characteristics of a sample of young adults residing in Puerto Rico, 2022-2025.

| Characteristics | n (%) |
| --- | --- |
| **Age** |  |
| mean ± SD | 27.3 ± 4.1 |
| 21 - 25 | 1076 (38.6) |
| 26 - 30 | 983 (35.32) |
| 31 - 35 | 724 (26.02) |
| **Area of residence** |  |
| Metropolitan area | 1310 (47.07) |
| Non-metropolitan area | 1473 (52.93) |
| **Level of Education** |  |
| Less than 12 years | 8 (0.29) |
| 12 years of GED | 315 (11.32) |
| 1 - 3 years of college | 793 (28.49) |
| ≥ 4 years of college | 1667 (59.9) |
| **Employment** |  |
| Employed full-time | 1720 (61.8) |
| Employed part-time | 519 (18.65) |
| Student | 295 (10.6) |
| Unemployed | 249 (9.94) |
| **Civil Status** |  |
| Married | 238 (8.6) |
| Divorced/Separated | 86 (3.1) |
| Single | 1608 (57.8) |
| Widowed | 6 (0.2) |
| Living with a partner | 399 (14.3) |
| Other | 446 (16) |
| Children |  |
| No | 2183 (78.4) |
| Yes | 600 (21.6) |

*Note.* SD = Standard Deviation

**Table 2.** Sex differences in health perception, preventive care indicators, and cardiometabolic risk factors in young adults residing in Puerto Rico, 2022-2025.

| Characteristics | Overall | Female | Male | *P* value |
| --- | --- | --- | --- | --- |
| n | 2,783 | 2,003 | 780 |  |
| Body Mass Index, n (%) |  |  |  |  |
| mean ± SD | 27.6 ± 7.3 | 27.6 ± 7.5 | 27.5 ± 6.6 | 0.82 |
| Underweight | 128 (4.60) | 100 (5) | 28 (3.6) |  |
| Healthy Weight | 1061 (38.12) | 774 (38.6) | 287 (36.8) |  |
| Overweight | 750 (26.95) | 519 (25.9) | 231 (29.6) |  |
| Obesity | 844 (30.33) | 610 (30.5) | 234 (30) |  |
| General Health Status, n (%) |  |  |  |  |
| Excellent | 851 (30.6) | 567 (28.3) | 284 (36.4) | < 0.001 |
| Very good | 1027 (36.9) | 737 (36.8) | 290 (37.2) |  |
| Good | 687 (24.7) | 524 (26.2) | 163 (20.9) |  |
| Regular | 206 (7.4) | 166 (8.3) | 40 (5.1) |  |
| Poor | 12 (0.4) | 9 (0.4) | 3 (0.4) |  |
| Poor Physical Health, n (%) |  |  |  |  |
| 0 - 5 days | 2214 (79.6) | 1571 (78.4) | 643 (82.4) | 0.01 |
| 6 - 10 days | 332 (11.9) | 256 (12.8) | 76 (9.7) |  |
| 11 - 14 days | 92 (3.3) | 61 (3) | 31 (4) |  |
| > 15 days | 145 (5.2) | 115 (5.7) | 30 (3.8) |  |
| Poor Mental Health, n (%) |  |  |  |  |
| 0 - 5 days | 1656 (59.5) | 1149 (57.4) | 507 (65) | < 0.001 |
| 6 - 10 days | 669 (24) | 510 (25.5) | 159 (20.4) |  |
| 11 - 14 days | 238 (8.6) | 187 (9.3) | 51 (6.5) |  |
| > 15 days | 220 (7.9) | 157 (7.8) | 63 (8.1) |  |
| Impairment Caused by Poor Health, n (%) |  |  |  |  |
| 0 - 5 days | 1960 (70.4) | 1376 (68.7) | 584 (74.9) | <0.01 |
| 6 - 10 days | 522 (18.8) | 399 (19.9) | 123 (15.8) |  |
| 11 - 14 days | 167 (6) | 134 (6.7) | 33 (4.2) |  |
| > 15 days | 134 (4.8) | 94 (4.7) | 40 (5.1) |  |
| Healthcare Insurance, n (%) |  |  |  |  |
| No | 269 (9.7) | 175 (8.7) | 94 (12.1) | 0.01 |
| Yes | 2514 (90.3) | 1828 (91.3) | 686 (87.9) |  |
| Afford Medical Attention in the Past Year, n (%) |  |  |  |  |
| No | 1756 (63.1) | 1255 (62.7) | 501 (64.2) | 0.44 |
| Yes | 1027 (36.9) | 748 (37.3) | 279 (35.8) |  |
| Have Primary Care Physician, n (%) |  |  |  |  |
| Yes, only one | 1574 (56.6) | 1169 (58.4) | 405 (51.9) | <0.01 |
| More than one | 311 (11.2) | 225 (11.2) | 86 (11) |  |
| No | 898 (32.3) | 609 (30.4) | 289 (37.1) |  |
| Time Since Last Physical Exam, n (%) |  |  |  |  |
| In the past year | 1748 (62.8) | 1338 (66.8) | 410 (52.6) | < 0.001 |
| In the past 2 years | 628 (22.6) | 434 (21.7) | 194 (24.9) |  |
| In the past 5 years | 238 (8.55) | 141 (7) | 97 (12.4) |  |
| > 5 years | 169 (6.07) | 90 (4.5) | 79 (10.1) |  |
| Time Since Last Dental Exam, n (%) |  |  |  |  |
| In the past year | 1791 (64.4) | 1315 (65.7) | 476 (61) | 0.01 |
| In the past 2 years | 599 (21.5) | 425 (21.2) | 174 (22.3) |  |
| In the past 5 years | 260 (9.3) | 183 (9.1) | 77 (9.9) |  |
| > 5 years | 133 (4.8) | 80 (4) | 53 (6.8) |  |
| Flu Shot in the Past Year, n (%) |  |  |  |  |
| No | 1938 (69.6) | 1405 (70.1) | 533 (68.3) | 0.35 |
| Yes | 845 (30.4) | 598 (29.9) | 247 (31.7) |  |
| PAP Smear Test, n (%) |  |  |  |  |
| No | 442 (22.1) | 442 (22.1) | n/a | n/a |
| Yes | 1561 (77.9) | 1561 (77.9) | n/a |  |
| HIV or STI Test, n (%) |  |  |  |  |
| No | 816 (29.3) | 453 (22.6) | 363 (46.5) | < 0.001 |
| Yes | 1967 (70.7) | 1550 (77.4) | 417 (53.5) |  |
| HPV vaccine, n (%) |  |  |  |  |
| No | 1261 (45.3) | 811 (40.5) | 450 (57.7) | < 0.001 |
| Yes | 1522 (54.7) | 1192 (59.5) | 330 (42.3) |  |
| High Blood Sugar/Diabetes Testing, n (%) |  |  |  |  |
| No | 959 (34.5) | 615 (30.7) | 344 (44.1) | < 0.001 |
| Yes | 1824 (65.5) | 1388 (69.3) | 436 (55.9) |  |
| Diagnosis of High Blood Sugar/Prediabetes, n (%) |  |  |  |  |
| No | 2375 (85.3) | 1693 (84.5) | 682 (87.4) | 0.94 |
| Yes | 408 (14.7) | 310 (15.5) | 98 (12.6) |  |
| Diagnosis of High Blood Pressure/Hypertension, n (%) |  |  |  |  |
| Borderline | 135 (4.9) | 82 (4.1) | 53 (6.8) | < 0.001 |
| No | 2344 (84.2) | 1723 (86) | 621 (79.6) |  |
| Yes | 304 (10.9) | 198 (9.9) | 106 (13.6) |  |
| Tobacco Use, n (%) |  |  |  |  |
| Some days | 114 (4.1) | 79 (3.9) | 35 (4.5) | < 0.001 |
| None | 2097 (75.4) | 1620 (80.9) | 477 (61.2) |  |
| Everyday | 42 (1.5) | 24 (1.2) | 18 (2.3) |  |
| No response | 530 (19) | 280 (14) | 250 (32.1) |  |
| Alcohol Use Frequency, n (%) |  |  |  |  |
| At least once a month | 827 (29.7) | 638 (31.9) | 189 (24.2) | < 0.001 |
| 2 - 4 times a month | 843 (30.3) | 641 (32) | 202 (25.9) |  |
| 2 - 3 times a week | 237 (8.5) | 161 (8) | 76 (9.7) |  |
| > 4 times a week | 30 (1.1) | 20 (1) | 10 (1.3) |  |
| Never | 400 (14.4) | 324 (16.2) | 76 (9.7) |  |
| No response | 446 (16) | 219 (10.9) | 227 (29.1) |  |

*Note.* SD = Standard Deviation

Regarding cardiometabolic health, a third reported never having been tested for high blood sugar or diabetes, and nearly 15% reported a diagnosis of prediabetes or elevated blood sugar. Around 11% of participants reported having been told by a doctor they had borderline or high blood pressure/hypertension. With respect to substance use, three-quarters reported no tobacco use. Most participants reported consuming alcohol less than 4 times a month, whereas 9.6% reported drinking alcohol at least twice a week (**Table 2**).

In terms of attitudes toward health behaviors (**Table 3**), most participants agreed or strongly agreed that engaging in regular physical activity is important for maintaining health, that a balanced diet supports good health, and that mental and emotional health are as important as physical health for overall well-being. Additionally, 97.3% reported interest in developing healthier habits to improve their lifestyle.

**Table 3.** Sex differences in loneliness and attitudes towards health in young adults residing in Puerto Rico, 2022-2025.

| Characteristics | Overall | Female | Male | *P* value |
| --- | --- | --- | --- | --- |
| n | 2,783 | 2,003 | 780 |  |
| Loneliness Scores, mean ± SD |  |  |  |  |
| Emotional | 1.5 ± 2 | 2.9 ± 2 | 2.8 ± 1.9 | 0.15 |
| Social | 1.3 ± 1 | 1.5 ± 1 | 1.4 ± 1 | 0.32 |
| Total | 2.9 ± 1.1 | 1.4 ± 1.1 | 1.3 ± 1.1 | 0.19 |
| Physical activity is important for maintaining health, n (%) |  |  |  |  |
| Strongly agree | 2146 (77.11) | 1522 (76) | 624 (80) | 0.12 |
| Agree | 552 (19.83) | 419 (20.9) | 133 (17.1) |  |
| Neutral | 75 (2.69) | 55 (2.7) | 20 (2.6) |  |
| Disagree | 3 (0.11) | 3 (0.1) | 0 (0) |  |
| Strongly agree | 7 (0.25) | 4 (0.2) | 3 (0.4) |  |
| A Balanced Diet is Important for Good Health, n (%) |  |  |  |  |
| Strongly agree | 2073 (74.49) | 1490 (74.4) | 583 (74.7) | 0.02 |
| Agree | 608 (21.85) | 449 (22.4) | 159 (20.4) |  |
| Neutral | 90 (3.23) | 60 (3) | 30 (3.8) |  |
| Disagree | 5 (0.18) | 1 (0) | 4 (0.5) |  |
| Strongly agree | 7 (0.25) | 3 (0.1) | 4 (0.5) |  |
| Mental and Emotional Health are as Important for Overall Well-being, n (%) |  |  |  |  |
| Strongly agree | 2435 (87.5) | 1771 (88.4) | 664 (85.1) | 0.01 |
| Agree | 301 (10.82) | 205 (10.2) | 96 (12.3) |  |
| Neutral | 36 (1.29) | 22 (1.1) | 14 (1.8) |  |
| Disagree | 3 (0.11) | (0) | 3 (0.4) |  |
| Strongly agree | 8 (0.29) | 5 (0.2) | 3 (0.4) |  |
| Interest in Developing Healthier Habits to Improve Lifestyle, n (%) |  |  |  |  |
| Strongly agree | 2285 (82.11) | 1672 (83.5) | 613 (78.6) | 0.01 |
| Agree | 422 (15.16) | 284 (14.2) | 138 (17.7) |  |
| Neutral | 63 (2.26) | 39 (1.9) | 24 (3.1) |  |
| Disagree | 2 (0.07) | (0) | 2 (0.3) |  |
| Strongly agree | 11 (0.4) | 8 (0.4) | 3 (0.4) |  |

*Note.* SD = Standard Deviation

We next examined sex differences across the study variables (**Tables 2**–**3**). There were no significant sex differences in mean BMI. However, compared to females, a statistically significantly greater proportion of males rated their general health as excellent, whereas a greater proportion of females rated their health as good or regular (χ²(4, 2783) = 26.00). With respect to physical health, a significantly higher proportion of females reported experiencing fewer than 11 days of poor physical health in the past month, whereas a higher proportion of males reported experiencing more than 14 days of poor physical health (χ²(3, 2783) = 10.81). In contrast, males were significantly more likely than females to report fewer than 11 days of poor mental health, while females were more likely to report 11 to 14 days of poor mental health in the past month (χ²(3, 2783) = 16.70). Similarly, most males reported that poor physical or mental health interfered with their daily activities for fewer than five days in the past month, whereas a significantly greater proportion of females reported interference lasting 6 to 14 days (χ²(3, 2783) = 14.08).

Additionally, a significantly greater proportion of males compared to females reported not having a PCP (χ²(2, 2783) = 11.82) or insurance (χ²(1, 2783) = 7.06). There were no sex differences in the ability to afford medical attention. Compared to males, a significantly greater proportion of females reported going to the dentist in the past year, while a greater proportion of males reported not having been to the dentist in 5 years or more (χ²(3, 2783) = 11.72). Similarly, significantly more females reported having a physical exam in the past year, whereas a greater proportion of males reported undergoing a physical exam more than 2 years ago (χ²(3, 2783) = 69.14). Females were significantly more likely than males to have been tested for HIV or other STDs (χ²(1, 2783) = 155.03) and to have received the HPV vaccine (χ²(1, 2783) = 67.05). There were no sex differences in receipt of the flu shot in the past year. With respect to cardiometabolic screening and diagnoses, a significantly greater proportion of females compared to males were tested for high blood sugar or diabetes (χ²(1, 2783) = 44.62). However, there were no significant sex differences in the diagnosis of high blood sugar or prediabetes. In contrast, a significantly greater proportion of males reported having been diagnosed with high or borderline blood pressure (χ²(2, 2783) = 18.23).

Regarding substance use, a statistically significantly greater proportion of males reported using tobacco than females (χ²(3, 2783) = 130.25) (**Table 2**). Further, while a significantly greater proportion of females denied consuming alcohol, they consumed alcohol more frequently (χ²(5, 2783) = 152.014).

While both groups rated regular physical activity as important for maintaining health similarly, more males disagreed that a balanced diet supports good health (χ²(4, 2783) = 11.963). Similarly, a significantly greater proportion of females strongly agreed that mental and emotional health are as important as physical health for overall well-being (χ²(4, 2783) = 13.090), and that they were interested in developing healthier habits to improve their lifestyle (χ²(4, 2783) = 14.507). There were no significant sex differences in the total loneliness score or its subscales.

Finally, we examined differences across three age groups (**Table 4**). There was a statistically significant association between age group and BMI category, with adults aged 21-25 being more likely to have a healthy weight, whereas adults aged 31-35 were less likely to have obesity, indicating increasing weight-related risk with age (χ²(6, 2783) = 59.95). There were no significant differences in perceived general health or the number of days of perceived poor physical health between age groups. However, poorer mental health was significantly more prevalent among younger adults, particularly those aged 21-25, whereas older participants (31–35) were more likely to report few or no days of poor mental health (χ²(6, 2783) = 51.195). Similarly, older adults (31-35) reported fewer days in which poor physical or mental health interfered with their daily activities compared to the younger groups (χ²(2, 2783) = 24.29).

**Table 4.** Age differences in health perception, preventive care indicators, and cardiometabolic risk factors in young adults residing in Puerto Rico, 2022-2025.

| Characteristics | 21-25 | 26-30 | 31-35 | *P* value |
| --- | --- | --- | --- | --- |
| n | 1,076 | 983 | 724 |  |
| Body Mass Index, n (%) |  |  |  |  |
| mean ± SD | 26.2 ± 6.6 | 28.1 ± 7.7 | 28.7 ± 7.2 | <0.001 |
| Underweight | 63<sup>a</sup> (5.9) | 42 (4.3) | 23<sup>b</sup> (3.2) |  |
| Healthy Weight | 489<sup>a</sup> (45.4) | 343<sup>b</sup> (34.9) | 229<sup>b</sup> (31.6) |  |
| Overweight | 260 (24.2) | 282 (28.7) | 208 (28.7) |  |
| Obesity | 264<sup>a</sup> (24.5) | 316<sup>b</sup> (32.1) | 264<sup>b</sup> (36.5) |  |
| General Health Status, n (%) |  |  |  |  |
| Excellent | 343 (31.9) | 285 (29) | 223 (30.8) | 0.46 |
| Very good | 395 (36.7) | 379 (38.6) | 253 (34.9) |  |
| Good | 253 (23.5) | 240 (24.4) | 194 (26.8) |  |
| Regular | 82 (7.6) | 72 (7.3) | 52 (7.2) |  |
| Poor | 3 (0.3) | 7 (0.7) | 2 (0.3) |  |
| Poor Physical Health, n (%) |  |  |  |  |
| 0 - 5 days | 832 (77.3) | 780 (79.3) | 602 (83.1) | 0.14 |
| 6 - 10 days | 144 (13.4) | 118 (12) | 70 (9.7) |  |
| 11 - 14 days | 40 (3.7) | 34 (3.5) | 18 (2.5) |  |
| > 15 days | 60 (5.6) | 51 (5.2) | 34 (4.7) |  |
| Poor Mental Health, n (%) |  |  |  |  |
| 0 - 5 days | 573<sup>a</sup> (53.3) | 584<sup>b</sup> (59.4) | 499<sup>c</sup> (68.9) | < 0.001 |
| 6 - 10 days | 309<sup>a</sup> (28.7) | 225<sup>b</sup> (22.9) | 135<sup>b</sup> (18.6) |  |
| 11 - 14 days | 111<sup>a</sup> (10.3) | 81 (8.2) | 46<sup>b</sup> (6.4) |  |
| > 15 days | 83 (7.7) | 93<sup>a</sup> (9.5) | 44<sup>b</sup> (6.1) |  |
| Impairment Caused by Poor Health, n (%) |  |  |  |  |
| 0 - 5 days | 705<sup>a</sup> (65.5) | 691<sup>a</sup> (70.3) | 564<sup>b</sup> (77.9) | < 0.001 |
| 6 - 10 days | 240<sup>a</sup> (22.3) | 175<sup>b</sup> (17.8) | 107<sup>b</sup> (14.8) |  |
| 11 - 14 days | 76<sup>a</sup> (7.1) | 65<sup>a</sup> (6.6) | 26<sup>b</sup> (3.6) |  |
| > 15 days | 55 (5.1) | 52 (5.3) | 27 (3.7) |  |
| Healthcare Insurance, n (%) |  |  |  |  |
| No | 70<sup>a</sup> (6.5) | 127<sup>b</sup> (12.9) | 72<sup>b</sup> (9.9) | < 0.001 |
| Yes | 1006<sup>a</sup> (93.5) | 856<sup>b</sup> (87.1) | 652<sup>b</sup> (90.1) |  |
| Afford Medical Attention in the Past Year, n (%) |  |  |  |  |
| No | 690 (64.1) | 604 (61.4) | 462 (63.8) | 0.41 |
| Yes | 386 (35.9) | 379 (38.6) | 262 (36.2) |  |
| Have Primary Care Physician, n (%) |  |  |  |  |
| Yes, only one | 617 (57.3) | 546 (55.5) | 411 (56.8) | 0.45 |
| More than one | 113 (10.5) | 106 (10.8) | 92 (12.7) |  |
| No | 346 (32.2) | 331 (33.7) | 221 (30.5) |  |
| Time Since Last Physical Exam, n (%) |  |  |  |  |
| In the past year | 625<sup>a</sup> (58.1) | 625<sup>b</sup> (63.6) | 498<sup>b</sup> (68.8) | <0.01 |
| In the past 2 years | 277 (25.7) | 215 (21.9) | 136 (18.8) |  |
| In the past 5 years | 102 (9.5) | 82 (8.3) | 54 (7.5) |  |
| > 5 years | 72 (6.7) | 61 (6.2) | 36 (5) |  |
| Time Since Last Dental Exam, n (%) |  |  |  |  |
| In the past year | 668<sup>a</sup> (62.1) | 612<sup>a</sup> (62.3) | 511<sup>b</sup> (70.6) | <0.01 |
| In the past 2 years | 256<sup>a</sup> (23.8) | 217 (22.1) | 126<sup>b</sup> (17.4) |  |
| In the past 5 years | 97 (9) | 107 (10.9) | 56 (7.7) |  |
| > 5 years | 55 (5.1) | 47 (4.8) | 31 (4.3) |  |
| Flu Shot in the Past Year, n (%) |  |  |  |  |
| No | 738 (68.6) | 686 (69.8) | 514 (71) | 0.55 |
| Yes | 338 (31.4) | 297 (30.2) | 210 (29) |  |
| PAP Smear Test, n (%) |  |  |  |  |
| No | 301<sup>a</sup> (40.6) | 105<sup>b</sup> (14.7) | 36<sup>c</sup> (6.6) | < 0.001 |
| Yes | 440<sup>a</sup> (59.4) | 611<sup>b</sup> (85.3) | 510<sup>c</sup> (93.4) |  |
| HIV or STI Test, n (%) |  |  |  |  |
| No | 457<sup>a</sup> (42.5) | 240<sup>b</sup> (24.4) | 119<sup>c</sup> (16.4) | < 0.001 |
| Yes | 619<sup>a</sup> (57.5) | 743<sup>b</sup> (75.6) | 605<sup>c</sup> (83.6) |  |
| HPV vaccine, n (%) |  |  |  |  |
| No | 385<sup>a</sup> (35.8) | 412<sup>b</sup> (41.9) | 464<sup>c</sup> (64.1) | < 0.001 |
| Yes | 691<sup>a</sup> (64.2) | 571<sup>b</sup> (58.1) | 260<sup>c</sup> (35.9) |  |
| High Blood Sugar/Diabetes Testing, n (%) |  |  |  |  |
| No | 424<sup>a</sup> (39.4) | 337<sup>b</sup> (34.3) | 198<sup>c</sup> (27.3) | < 0.001 |
| Yes | 652<sup>a</sup> (60.6) | 646<sup>b</sup> (65.7) | 526<sup>c</sup> (72.7) |  |

| Diagnosis of High Blood Sugar/Prediabetes, n (%) |  |  |  |  |
| --- | --- | --- | --- | --- |
| No | 929 (86.3) | 839 (85.4) | 607 (83.8) | 0.48 |
| Yes | 147 (13.7) | 144 (14.6) | 117 (16.2) |  |
| Diagnosis of High Blood Pressure/Hypertension, n (%) |  |  |  |  |
| Borderline | 31<sup>a</sup> (2.9) | 64<sup>b</sup> (6.5) | 40<sup>c</sup> (5.5) | < 0.001 |
| No | 974<sup>a</sup> (90.5) | 810<sup>b</sup> (82.4) | 560<sup>c</sup> (77.3) |  |
| Yes | 71<sup>a</sup> (6.6) | 109<sup>b</sup> (11.1) | 124<sup>b</sup> (17.1) |  |
| Tobacco Use, n (%) |  |  |  |  |
| Some days | 47 (4.4) | 44 (4.5) | 23 (3.2) | < 0.001 |
| None | 765<sup>a</sup> (71.1) | 766<sup>b</sup> (77.9) | 566<sup>b</sup> (78.2) |  |
| Everyday | 15 (1.4) | 13 (1.3) | 14 (1.9) |  |
| No response | 249<sup>a</sup> (23.1) | 160<sup>b</sup> (16.3) | 121<sup>b</sup> (16.7) |  |
| Alcohol Use Frequency, n (%) |  |  |  |  |
| At least once a month | 313 (29.1) | 299 (30.4) | 215 (29.7) | <0.01 |
| 2 - 3 times a week | 93 (8.6) | 89 (9.1) | 55 (7.6) |  |
| 2 - 4 times a month | 301<sup>a</sup> (28) | 297 (30.2) | 245<sup>b</sup> (33.8) |  |
| > 4 times a week | 8 (0.7) | 14 (1.4) | 8 (1.1) |  |
| Never | 147 (13.7) | 150 (15.3) | 103 (14.2) |  |
| No response | 214<sup>a</sup> (19.9) | 134<sup>b</sup> (13.6) | 98<sup>b</sup> (13.5) |  |

*Notes.* SD = Standard Deviation; Values with different superscript letters (a, b, c) indicate statistically significant differences between groups after correcting for multiple comparisons.

Regarding access to and use of healthcare services, adults aged 21-25 were more likely to have insurance than other age groups (χ²(2, 2783) = 24.29). There were no significant group differences in the frequency of visits to a PCP or in the ability to afford medical attention. Older participants (31–35) reported more recent dental care utilization, whereas younger adults (21–25) were more likely to report longer intervals since their last dental visit, particularly in the 1-2 years range (χ²(6, 2783) = 19.570). The same pattern was observed for the time since their last physical exam (χ²(6, 2783) = 21.939). Further, the number of people who have been tested for HIV or other STDs increased with age (χ²(2, 2783) =159.212), while the number of people having received the HPV vaccine decreased with age (χ²(6, 2783) = 147.037).

With respect to cardiometabolic factors, the older the group, the more likely to have been tested for high blood sugar or diabetes (χ²(2, 2783) = 27.879). There were no age differences in the number of people who reported receiving a diagnosis of prediabetes or high blood sugar *(P =* .481). In contrast, there was a statistically significant association between age group and diagnosed high blood pressure or hypertension (χ²(4, 2783) = 68.342), with prevalence increasing across age groups, from 6.6% in ages 21-25 to 17.1% in ages 31-35.

In terms of substance use, the older the group, the higher the prevalence of tobacco use (χ²(6, 2783) = 22.941). There was also a significant association between age group and alcohol use, with adults aged 31-35 being more likely to report drinking 2 to 4 times per month (χ²(10, 2783) = 25.773), while other drinking frequencies did not differ meaningfully by age.

There were no significant age differences in the level of agreement about whether regular physical activity is important for maintaining health, a balanced diet for supporting good health, considering that mental and emotional health are as important as physical health for overall well-being, or interest in developing healthier habits to improve their lifestyle.

Regarding loneliness, there was a statistically significant difference in total loneliness score between age groups (F(2, 2334) = 4.25, *P* = .01, η² = .004) (**Table 5**). Participants aged 21-25 reported significantly higher total loneliness than those aged 31-35, whereas no significant differences were observed with the 26-30 age group. Further, emotional loneliness differed by age group (F(2, 2334) = 7.71, *P* < .001, η² = .007), with those aged 21-25 reporting significantly higher emotional loneliness than those aged 26-30 and 31-35, while the latter two groups did not differ from each other. There were no significant age group differences on the social loneliness scale.

**Table 5.** Age differences in loneliness and attitudes towards health in young adults residing in Puerto Rico, 2022-2025.

| Characteristics | 21-25<br>n = 1,076 | 26-30<br>n = 983 | 31-35<br>n = 724 | P value |
| --- | --- | --- | --- | --- |
| Loneliness Scores, mean ± SD |  |  |  |  |
| Emotional | 1.59<sup>a</sup> ± 1 | 1.45<sup>b</sup> ± 1 | 1.39<sup>b</sup> ± 1 | < 0.001 |
| Social | 1.48 ± 1.1 | 1.51 ± 1.1 | 1.40 ± 1.2 | 0.086 |
| Total | 3.13<sup>a</sup> ± 2 | 3.01 ± 2 | 2.85<sup>b</sup> ± 2.1 | 0.014 |
| Physical activity is important for maintaining health, n (%) |  |  |  |  |
| Strongly agree | 819 (76.1) | 761 (77.4) | 566 (78.2) | 0.786 |
| Agree | 218 (20.3) | 191 (19.4) | 143 (19.8) |  |
| Neutral | 34 (3.2) | 28 (2.8) | 13 (1.8) |  |
| Disagree | 1 (0.1) | 1 (0.1) | 1 (0.1) |  |
| Strongly agree | 4 (0.4) | 2 (0.2) | 1 (0.1) |  |
| A Balanced Diet is Important for Good Health, n (%) |  |  |  |  |
| Strongly agree | 789 (73.3) | 733 (74.6) | 551 (76.1) | 0.440 |
| Agree | 239 (22.2) | 213 (21.7) | 156 (21.5) |  |
| Neutral | 44 (4.1) | 31 (3.2) | 15 (2.1) |  |
| Disagree | 1 (0.1) | 3 (0.3) | 1 (0.1) |  |
| Strongly agree | 3 (0.3) | 3 (0.3) | 1 (0.1) |  |
| Mental and Emotional Health are as Important for Overall Well-being, n (%) |  |  |  |  |
| Strongly agree | 952 (88.5) | 852 (86.7) | 631 (87.2) | 0.619 |
| Agree | 102 (9.5) | 114 (11.6) | 85 (11.7) |  |
| Neutral | 16 (1.5) | 13 (1.3) | 7 (1) |  |
| Disagree | 2 (0.2) | 1 (0.1) | (0) |  |
| Strongly agree | 4 (0.4) | 3 (0.3) | 1 (0.1) |  |
| Interest in Developing Healthier Habits to Improve Lifestyle, n (%) |  |  |  |  |
| Strongly agree | 907 (84.3) | 800 (81.4) | 578 (79.8) | 0.169 |
| Agree | 137 (12.7) | 158 (16.1) | 127 (17.5) |  |
| Neutral | 25 (2.3) | 20 (2) | 18 (2.5) |  |
| Disagree | 1 (0.1) | 1 (0.1) | (0) |  |
| Strongly agree | 6 (0.6) | 4 (0.4) | 1 (0.1) |  |

*Notes.* SD = Standard Deviation; Values with different superscript letters (a, b, c) indicate statistically significant differences between groups after correcting for multiple comparisons.

## DISCUSSION

This study offers the first post-pandemic, multi-domain characterization of health risk among young adults in Puerto Rico, a population that has experienced an unprecedented sequence of compounding crises during a developmentally critical period. Our findings reveal a high prevalence of self-reported adverse health indicators associated with elevated risk for chronic disease and poor health outcomes. Notably, younger groups were less likely to report tobacco or alcohol use, behaviors strongly associated with elevated long-term risk of disease^22–25^, suggesting a potential window for reinforcing protective behaviors early in adulthood. Despite this, substantial gaps in preventive care were observed. Approximately one-third of participants reported not having a PCP and not having visited a PCP or dentist in the past year, with this pattern more common among males. Influenza vaccination uptake was low, and over half of the participants were classified as having overweight or obesity. The observed obesity prevalence (30.3%) is comparable to NHANES estimates for adults aged 20–39 (35.5%)^26^, with the modest difference likely attributable to age-group categorization, underscoring the concerning burden of obesity in this age group and its downstream implications across the lifespan.

Cardiometabolic risk factors were also prevalent. Nearly 11% reported a diagnosis of high blood pressure, three times higher among those aged 31–35 than those aged 21–25, and males were more likely than females to report borderline or high blood pressure. These self-reported estimates are likely to substantially undercount the true prevalence, as nationally, only one in four young adults with hypertension is aware of their diagnosis, suggesting the burden in this population may be substantially higher than captured in this study^27^. Similarly, 14.7% of participants reported high blood glucose levels, exceeding national estimates for diabetes prevalence in this age group. These findings align with prior evidence of elevated cardiometabolic burden among Puerto Ricans^1,2,4,6,7,16^, and are especially concerning given that adverse cardiometabolic profiles in early adulthood predict long-term outcomes, including cardiovascular disease, chronic kidney disease, cancer, and neurodegenerative disease^8–10^, as well as substantial economic costs through increased healthcare utilization and lost productivity^28–30^.

Despite high insurance coverage in this study population, over 36% of young adults reported difficulty affording medical care in the past year, exceeding rates reported in the BRFSS and national affordability studies ^20,31^. In Puerto Rico, where Medicaid reimbursement is capped well below rates in US states, nominal coverage does not translate into adequate access, particularly for young adults who have aged out of pediatric care without a clear pathway into adult preventive services. Targeted strategies, including sliding-scale clinics, university-based health services, and community health worker programs, are needed to bridge the gap between enrollment and meaningful care engagement.

Mental health challenges were also prominent. Over one-third reported at least one week of poor mental health in the past month, which often interfered with their usual activities. Females reported greater mental health burden and functional interference than males^32–34^. Loneliness scores exceeded those reported in comparable US and international samples^35–37^, with younger adults (21–25) reporting the highest levels, consistent with evidence that loneliness peaks in young adulthood^38^. Given well-established links between loneliness and diabetes, obesity, depression, suicidal ideation, substance use, and reduced preventive care engagement^35,39,40^, integrating mental health screening into primary and community care settings should be a priority. The elevated loneliness observed may also reflect disruptions of social networks following the pandemic.

Encouragingly, participants expressed strongly positive attitudes toward physical activity, nutrition, and mental health. The disconnect between these favorable attitudes and the adverse profiles observed points to structural barriers, not motivation, as the primary impediment to healthy behavior. Interventions should therefore prioritize removing access barriers and embedding health resources within settings young adults already frequent, rather than focusing on attitude change.

Limitations include reliance on self-report, which increases recall and desirability bias, and recruitment at metropolitan concerts and sporting events, which may overrepresent urban young adults and underestimate burden in rural or more isolated subgroups. Information on diet, income, and substance use was not collected. The cross-sectional design precludes causal inference, and longitudinal studies are needed to establish the temporal ordering of exposures and outcomes.

Strengths include a large sample with power to detect subgroup differences, use of validated instruments adapted from the BRFSS, enabling national comparisons, contemporary post-pandemic data collection, and inclusion of participants from both rural and urban areas of Puerto Rico.

Taken together, these findings document a substantial and intersecting burden of cardiometabolic risk, psychosocial adversity, and preventive care gaps among young adults in Puerto Rico, a population that has navigated economic crisis, natural disaster, and pandemic within a single developmental decade. The coexistence of these risks alongside strong health motivation underscores that barriers are structural, not attitudinal, and that the window for intervention remains open. Integrated, multi-level strategies that expand affordable preventive services, embed mental health and cardiometabolic screening in community settings, and leverage cultural strengths are needed. Investments in this population, through sustained funding and policies that address Puerto Rico’s structural disadvantages within the US health system, hold the potential to reduce decades of preventable disease burden and narrow persistent health disparities in Puerto Rican and broader Latino communities.

## Data Availability

All data produced in the present study are available upon reasonable request to Varmed Management Group LLC.

## ACKNOWLEDGEMENTS

We thank all participants for contributing their time to this study. We also thank Lay Wah Fong and Andrés Martínez from Varmed Management Group LLC’s Data Analytics Department for their assistance in preparing the dataset for the study, and Karla M Andreu for providing feedback on the manuscript.

## DISCLAIMER

Dr. Guzmán-Vélez and Dr. Caballero are paid consultants for Melody Matters: A Health Project. Marisela Irizarry is employed by Varmed Management. Ms. Beaumont has no conflicts of interest to disclose.

